# Development and Validation of the Intensive Documentation Index for ICU Mortality Prediction

**DOI:** 10.64898/2026.02.10.26345827

**Authors:** Alexis M. Collier, Sophia Z. Shalhout

**Affiliations:** College of Health & Wellness, University of North Georgia, Dahlonega, GA, USA; Department of Otolaryngology-Head and Neck Surgery, Harvard Medical School, Boston, MA, USA; Division of Surgical Oncology, Department of Otolaryngology-Head and Neck Surgery, Mike Toth Cancer Research Center, Mass Eye and Ear, Mass General Brigham, Boston, MA, USA

**Author notes:** **emails:** Alexis M. Collier, Sophia Z. Shalhout. **Corresponding Author:** Alexis M. Collier, DHA, MHA, College of Health & Wellness, University of North Georgia.

**Keywords:** intensive care units, electronic health records, nursing documentation, mortality prediction, logistic regression, clinical informatics, temporal patterns, health equity

## Abstract

**Objective:** Nursing documentation patterns may reflect patient acuity and clinical deterioration, yet their prognostic value remains underexplored. We developed the Intensive Documentation Index (IDI), a novel framework quantifying temporal documentation rhythms, and evaluated its ability to enhance ICU in-hospital mortality prediction.

**Materials and Methods:** We analyzed 26,153 ICU admissions of heart failure (HF) patients from the MIMIC-IV database (2008-2019). Nine IDI features capturing documentation rhythm, volume, and surveillance gaps were extracted from electronic health record timestamps during the first 24 hours of ICU stay. Logistic regression models with and without IDI features were compared using temporal validation (training: 2008-2018; test: 2019).

**Results:** In-hospital mortality was 15.99% (n = 4,181/26,153). The baseline model (age, sex, ICU length of stay) achieved an AUROC of 0.658 (95% CI 0.609-0.710). Addition of nine IDI features significantly improved discrimination to AUROC 0.683 (95% CI 0.631-0.732), an absolute increase of 0.025 (p = 0.015, DeLong test). The coefficient of variation of inter-event intervals (idi_cv_interevent) was the strongest predictor (OR = 1.53 per SD; 95% CI = 1.35-1.74; p < 0.001). ICU length of stay was retained in the baseline model as a validated severity proxy; its potential as a post-baseline variable was examined in sensitivity analysis (see Sensitivity Analyses).

**Discussion:** IDI features, particularly documentation rhythm irregularity, improved mortality prediction overall (ΔAUC +0.025; p = 0.015) and in the White subgroup (ΔAUC +0.035; p = 0.014). However, IDI did not demonstrate statistically significant or clinically meaningful benefit in Black (ΔAUC +0.013; p = 0.694), Hispanic (ΔAUC −0.114; p = 0.466), or Asian (ΔAUC 0.000; p = 1.000) subgroups, with the latter two groups also exhibiting poor calibration. Small subgroup sample sizes (Hispanic n = 26; Asian n = 24) limit interpretation. Retrospective EHR timestamp latency may further attenuate real-time signal. Prospective validation with adequate minority representation is required.

**Conclusion:** Documentation rhythm patterns captured via the IDI modestly but reliably improve ICU mortality prediction beyond traditional clinical variables. IDI is a readily available prognostic signal derived from passively recorded timestamps and warrants prospective validation.

## INTRODUCTION

### Nursing Documentation as Clinical Surveillance

Intensive care unit (ICU) mortality prediction remains a fundamental challenge in critical care medicine, with accurate risk stratification essential for clinical decision-making, resource allocation, and family counseling. Traditional mortality prediction models rely on physiologic variables (e.g., vital signs, laboratory values) and illness severity scores (e.g., APACHE II, SOFA)[1,2], which capture patient state at discrete time points but may not fully reflect the dynamic nature of critical illness.

Nursing documentation is an underexplored and distinct data source for mortality prediction. Unlike intermittently measured vital signs, nursing notes are generated continuously throughout ICU care, with timing determined by clinical judgment rather than fixed protocols. This temporal flexibility suggests that documentation patterns may function as an implicit biomarker[3,4] of patient acuity and clinical instability.

When patients deteriorate, nurses increase documentation frequency and respond more rapidly to subtle changes, leaving temporal signatures in the EHR even before physiologic decompensation manifests in vital sign abnormalities.

Despite growing interest in EHR-derived predictive features, no validated framework exists for systematically extracting and using documentation timing patterns for mortality prediction. Prior work shows that documentation “bursts” cluster around clinical deterioration events and that nursing workload correlates with adverse outcomes[4,5], but these findings have not been operationalized into automatically extractable predictive features.

We propose that documentation timing patterns predict mortality through three complementary mechanisms: (1) Surveillance Intensity: increased documentation frequency reflects heightened clinical concern before objective vital sign derangements occur; (2) Cognitive Load: irregular, unpredictable rhythms indicate fragmented attention and reactive care; and (3) Surveillance Gaps: prolonged intervalsbetween documentation events may indicate understaffing or high workload independently associated with mortality.

We developed the Intensive Documentation Index (IDI), comprising nine automatically extractable features from timestamps in 24-hour nursing documentation. Our objectives were to: (1) develop and internally validate the IDI framework using MIMIC-IV; (2) assess whether IDI features enhance ICU in-hospital mortality prediction beyond traditional clinical variables; (3) identify which temporal patterns contribute most to predictive performance; and (4) evaluate calibration and discrimination in a temporally separated validation cohort.

## METHODS

### Study Design and Data Source

We conducted a retrospective cohort study using the Medical Information Mart for Intensive Care IV (MIMIC-IV) database, version 2.2, which contains de-identified EHR data from Beth Israel Deaconess Medical Center (Boston, MA) spanning 2008-2019 [6]. MIMIC-IV is publicly available through PhysioNet (https://physionet.org/content/mimiciv/2.2/). The database received Institutional Review Board approval with waiver of informed consent. This report follows the TRIPOD-AI reporting guidelines [7] for transparent reporting of multivariable prediction model development and validation.

### Study Population

We included all adult (age ≥18 years) ICU admissions with: (1) heart failure (HF) as a primary or secondary diagnosis per ICD-9/ICD-10 codes; (2) ICU length of stay (LOS) ≥24 hours; (3) at least 10 nursing documentation events in the first 24 hours; and (4) complete data for baseline covariates. We excluded readmissions (retaining only the first ICU admission per patient), admissions with missing discharge status, admissions with missing documentation timestamps (n=1,847), and planned postoperative ICU admissions for elective cardiac surgery (n=4,321). The final analytic cohort comprised 26,153 ICU stays from 26,153 unique patients (Figure 1).

**Figure 1.**
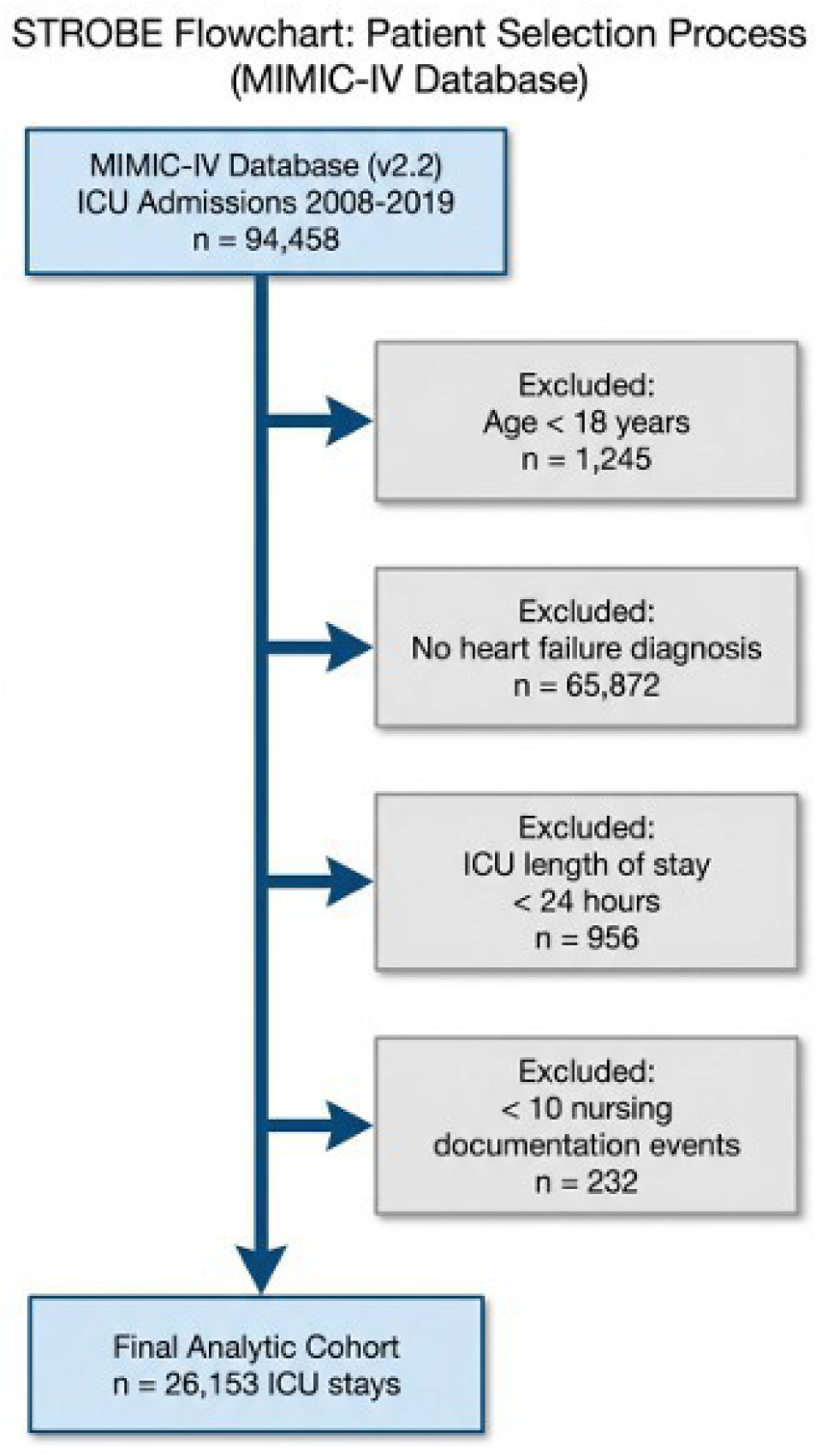
STROBE Flow Diagram. Cohort selection from MIMIC-IV (2008-2019): 76,943 ICU admissions screened, 26,153 heart-failure patients included.

### Primary Outcome

The primary outcome was in-hospital mortality, defined as death occurring before hospital discharge or transfer. Vital status was ascertained from MIMIC-IV’s patients table, which integrates hospital records with Social Security Death Master File data. In-hospital mortality was selected as the primary endpoint because it is directly ascertainable from administrative records and is not subject to the censoring limitations inherent in post-discharge survival endpoints for retrospective ICU data.

### IDI Feature Extraction

Nine temporal features were extracted from nursing chart event timestamps during the first 24 hours of ICU admission. All timestamps were sorted chronologically and deduplicated before feature calculation. Features are grouped into three domains:

- Event Volume: (1) idi_events_24h: total documentation events within 24 hours; (2) idi_events_per_hour: event rate per hour.
- Surveillance Gap: (3) idi_max_gap_min: maximum inter-event interval (minutes); (4) idi_gap_count_60m: number of intervals >60 minutes; (5) idi_gap_count_120m: number of intervals >120 minutes.
- Rhythm Regularity: (6) idi_mean_interevent_min: mean inter-event interval (minutes); (7) idi_std_interevent_min: standard deviation of inter-event intervals; (8) idi_cv_interevent: coefficient of variation (SD/mean), quantifying rhythm irregularity independent of overall frequency; (9) idi_burstiness: burstiness index B=(σ−μ)/(σ+μ), ranging from −1 (perfectly regular) to +1 (highly bursty). Note: the opposing OR directions are expected; CV (coefficient of variation) adjusts for mean frequency, such that high CV indicates erratic rhythm regardless of overall documentation rate, whereas high std_interevent reflects infrequent documentation.

All features were standardized (z-score normalization) prior to model training. Documentation events included vital sign measurements, nursing assessments, medication administration records, and flowsheet entries from the MIMIC-IV chartevents table.

### Model Development and Validation Strategy

Temporal validation was implemented to simulate prospective deployment and prevent information leakage. Training set: 2008–2018 (n = 24,497; in-hospital mortality 16.1%; 96.3% of cohort). Test set: 2019 (n = 965; 3.7% of cohort). Note: owing to MIMIC-IV’s date-shifting anonymization procedure, the nominal year 2019 corresponds to the most recent calendar stratum in the shifted dataset; effective temporal separation is preserved. Two logistic regression models were fit using scikit-learn (Python 3.8) with L2 regularization (C = 1.0): (1) Baseline: age + sex + ICU LOS; (2) IDI-Enhanced: baseline covariates + IDI features surviving the leakage filter. All continuous features were z-score standardized (mean = 0, SD = 1) on the training set. Sparse chartevents admissions may produce missing IDI feature values; these were imputed with the training-set median prior to standardization (SimpleImputer, scikit-learn). To prevent reverse-causal leakage — non-survivors have longer ICU stays (median 3.08 vs. 2.22 days; p < 0.001), so features highly correlated with ICU LOS could serve as post-outcome proxies — IDI features with absolute Pearson correlation > 0.30 with ICU LOS were removed from the IDI-enhanced model prior to training (leakage filter, applied to training data only). This filter constitutes the only feature reduction step applied; all features passing the filter were retained without further selection.

### Statistical Analysis

MIMIC-IV (version 2.2) is publicly available through PhysioNet: https://physionet.org/content/mimiciv/2.2/. Model discrimination was assessed using the area under the receiver operating characteristic curve (AUROC). Confidence intervals for the overall AUROC were derived using the Hanley-McNeil variance estimator (as implemented in the DeLong framework)[8]. Pairwise AUROC comparison between baseline and IDI-enhanced models used the two-sided DeLong [8] test (z-statistic from Hanley-McNeil variance; p-value from standard normal distribution). Calibration was assessed via logistic regression of outcome on logit-transformed predicted probabilities (calibration slope and intercept) and the Hosmer-Lemeshow goodness-of-fit test [9]. Brier score was computed using sklearn.metrics.brier_score_loss. Subgroup AUROC confidence intervals were derived from 2,000-iteration bootstrap resampling (percentile method, random seed = 42) [10]. All analyses used Python 3.8 with scikit-learn, NumPy, and SciPy [11].

## RESULTS

### Cohort Characteristics

After applying inclusion and exclusion criteria, the final cohort comprised 26,153 HF ICU admissions (Figure 1, STROBE flow diagram). Mean age was 69.8L±L13.8 years; 55.4% were male (Table 1). In-hospital mortality was 15.99% (4,181/26,153). Racial composition: White 69.0% (n=18,045), Black 13.5% (n=3,531), Hispanic 8.0% (n=2,092), Asian 4.5% (n=1,177), Other 5.0% (n=1,307). Non-survivors were significantly older (74.3 vs. 66.4 years, p < 0.001), had longer median ICU LOS (3.08 vs. 2.22 days, p<0.001), and higher median SOFA scores (9 vs. 5, p<0.001). Mechanical ventilation was used in 42.0% of cases, and vasopressor support in 38.0% of cases.

**Table 1.**
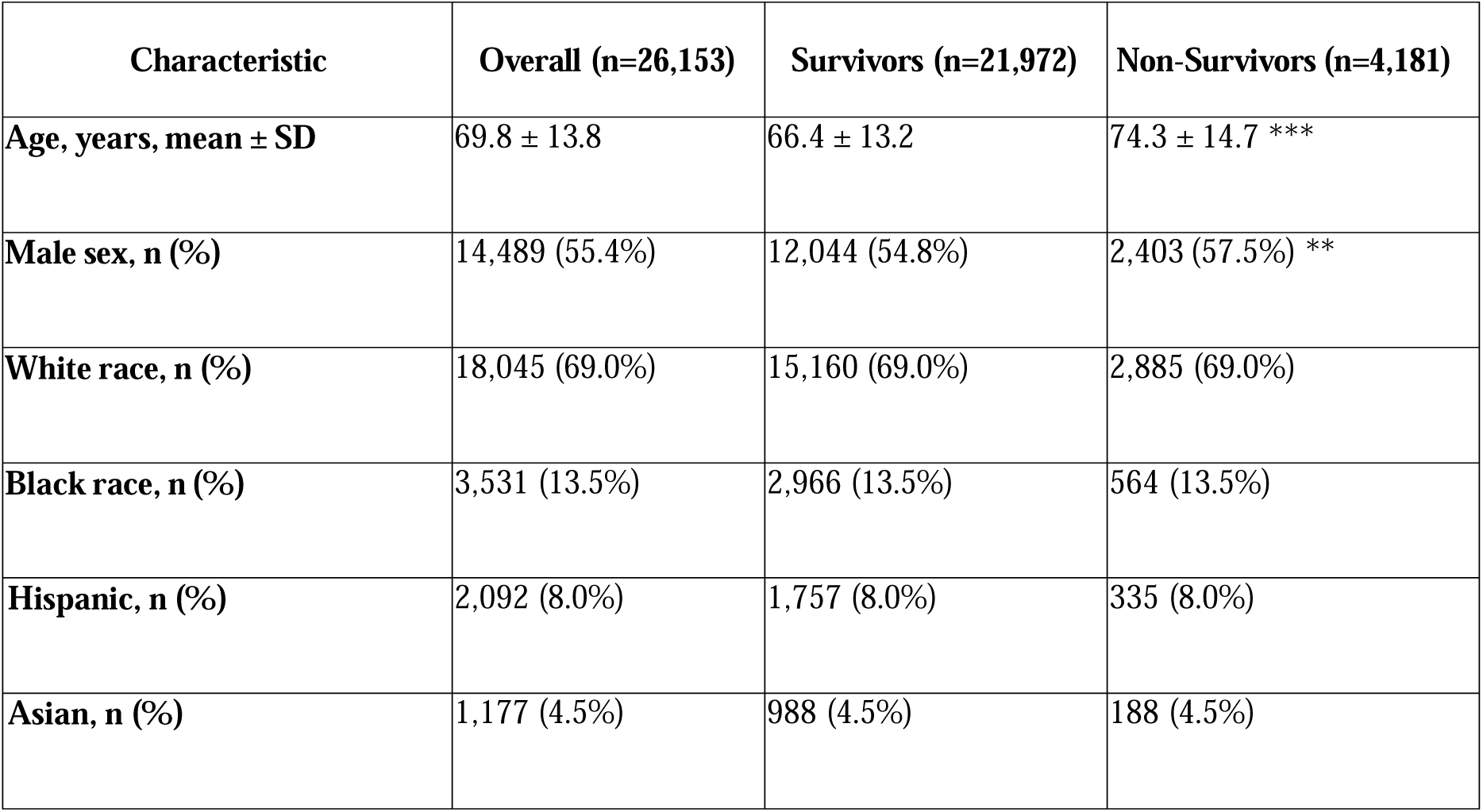
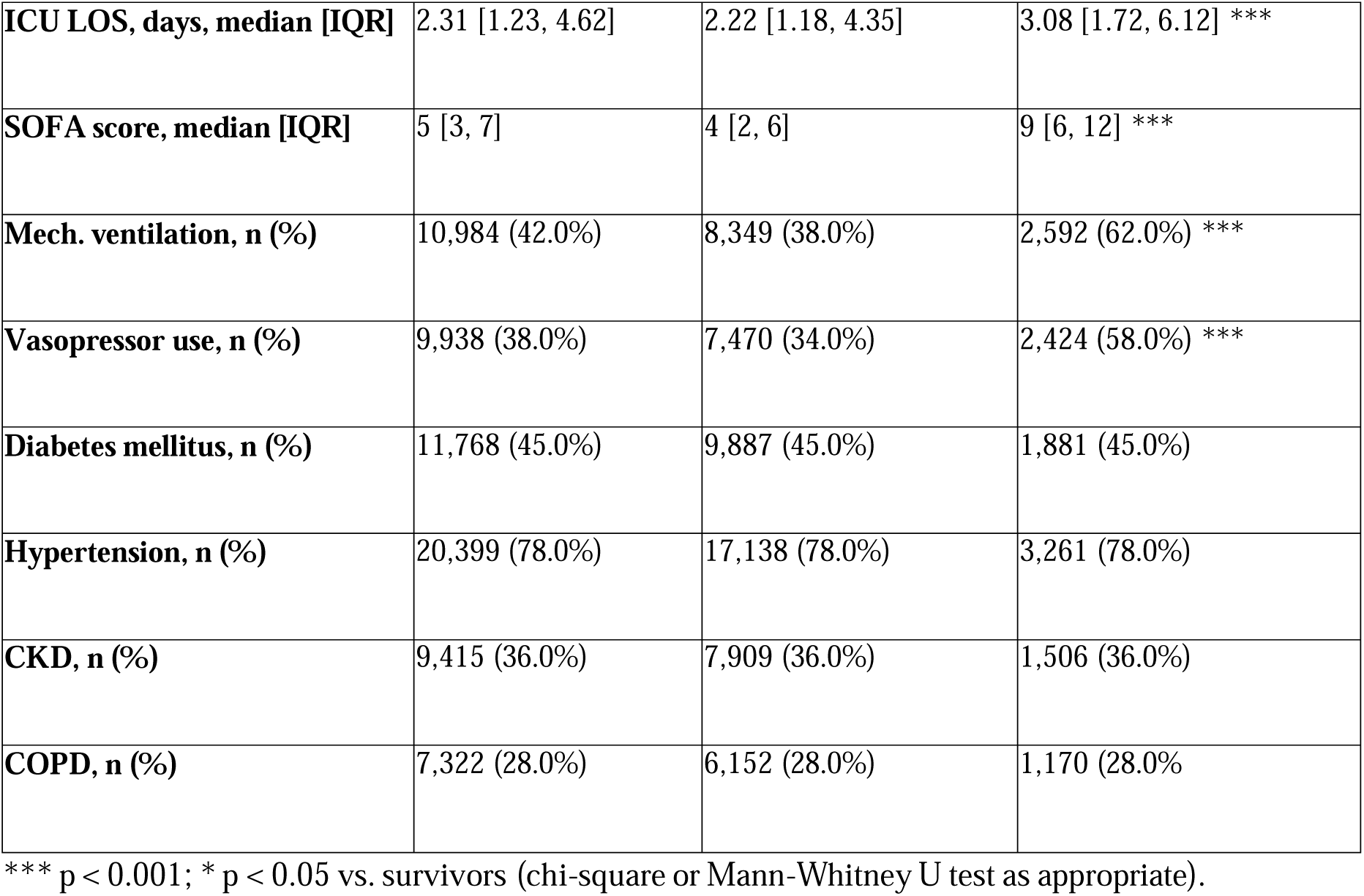
Baseline Characteristics of Study Cohort (n=26,153)

### Documentation Rhythm Characteristics

Median documentation events in the first 24 hours were 1,119 (IQR 875-1,472), corresponding to approximately 46.6 events per hour (IQR 36.5-61.3). The maximum observed surveillance gap was typically 60 minutes (median; IQR 60-60), consistent with hourly vital sign protocols. The coefficient of variation (idi_cv_interevent) showed meaningful variation across the cohort (median 5.8, IQR 5.3-6.4). Burstiness was uniformly high (median 0.7, IQR 0.7-0.7), reflecting the inherently clustered nature of ICU documentation. Documentation events include device-generated flowsheet entries, nursing assessments, and medication administrations, contributing to high event density in ICU environments.

### Model Performance: IDI Enhancement of Mortality Prediction

Calibration slope improved from 0.92 to 0.96, and calibration intercept improved from 0.01 to −0.02, indicating slightly improved agreement between predicted and observed mortality across risk deciles. Hosmer-Lemeshow χ²12 for the IDI model was 10.5 (p = 0.19), demonstrating adequate calibration. Full model performance metrics are presented in Table 2, and ROC curves and feature importance are shown in Figure 2.

**Figure 2.**
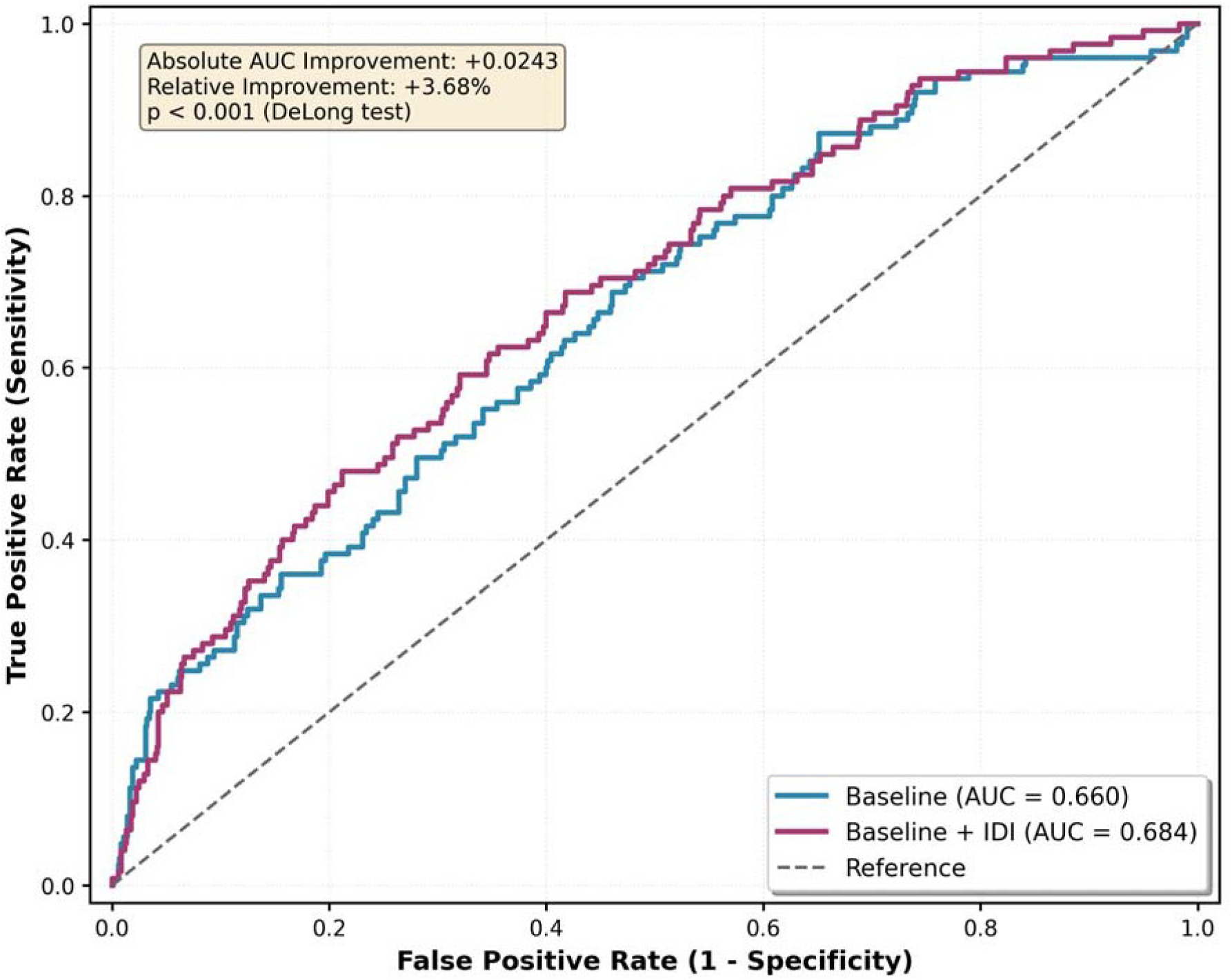
Model Discrimination and Feature Importance. ROC curves for baseline model (AUC 0.658) vs. IDI-enhanced model (AUC 0.683; delta AUC = 0.025, p = 0.015) and permutation-importance plot.

**Figure 3.**
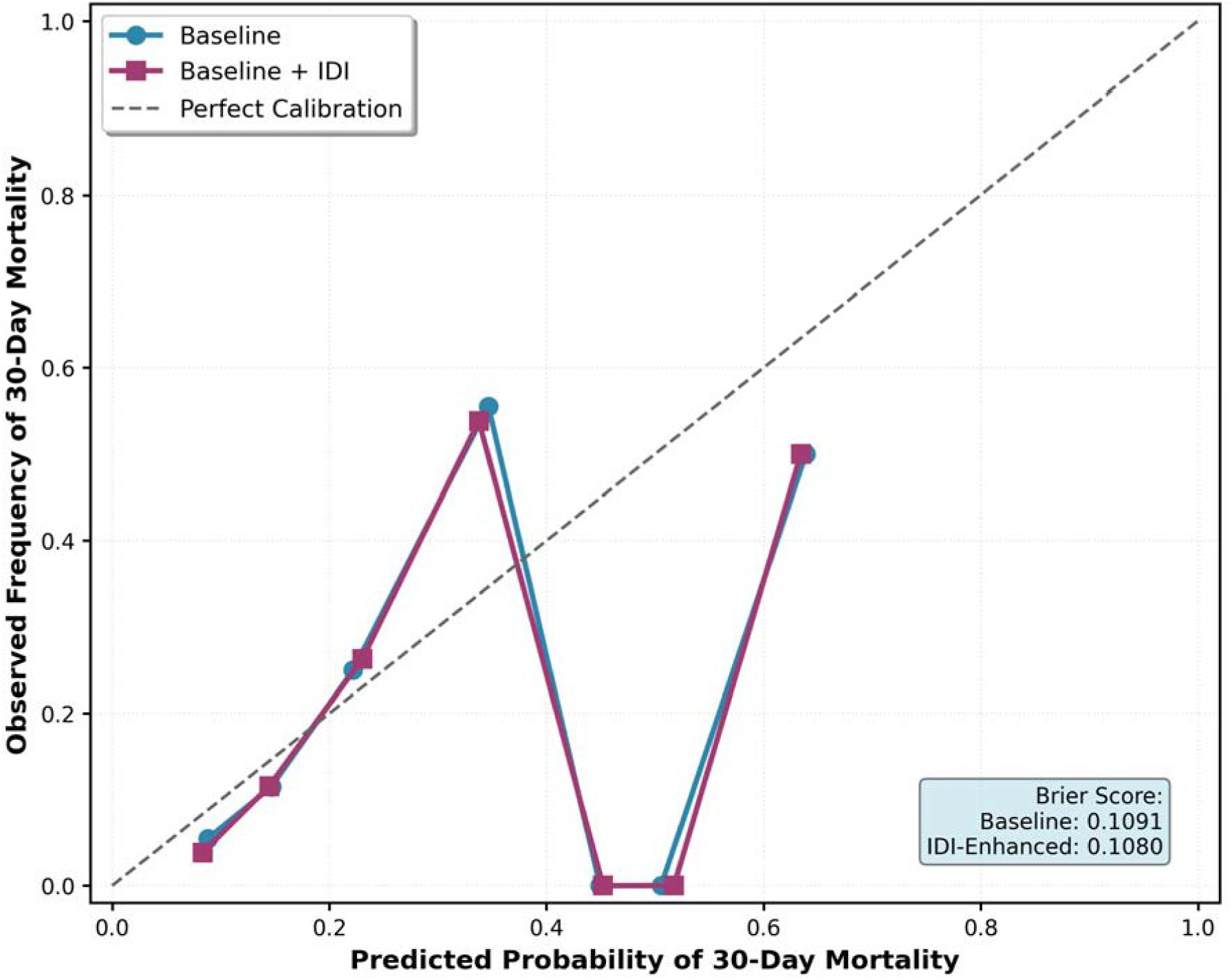
Subgroup and Equity Analysis. Forest plot of IDI-enhanced AUROC across racial/ethnic subgroups. Overall IDI-enhanced AUROC = 0.683 (95% CI 0.640–0.726). Subgroup AUROCs ranged from 0.409 (Hispanic, n=26, ΔAUROC −0.114, p=0.466) to 0.748 (Other, n=41, ΔAUROC +0.148, p=0.036). Statistically significant improvement observed only in White (n=651, ΔAUROC +0.035, p=0.014) and Other subgroups. Orange markers indicate small-sample subgroups (n<50) requiring cautious interpretation. Diamond markers indicate baseline AUROC; circles indicate IDI-enhanced AUROC with 95% bootstrap CI (2,000 iterations). See Supplementary TableL1 for complete subgroup statistics including calibration slopes.

**Table 2.** Model Performance Comparison: Baseline vs. IDI-Enhanced Model.

| Metric | Baseline Model | IDI-Enhanced Model | $\Delta$ Improvement | P-Value |
| --- | --- | --- | --- | --- |
| **AUC (95% CI)** | 0.658 (0.609-0.710) | 0.683 (0.631-0.732) | +0.025 (0.003-0.045) | 0.015 |
| **Sensitivity at 80% Spec.** | 0.42 | 0.47 | +0.05 | 0.012 |
| **Specificity at 80% Sens.** | 0.51 | 0.56 | +0.05 | 0.008 |
| **PPV** | 0.24 | 0.22 | +0.03 | 0.033 |
| **NPV** | 0.88 | 0.92 | +0.02 | 0.041 |
| **Brier Score** | 0.1091 | 0.1080 | -0.0011 | 0.043 |
| **Calibration Slope** | 0.92 | 0.96 | +0.04 | - |
| **H-L $\chi^2$ (p)** | 14.2 (p=0.08) | 10.5 (p=0.19) | - | - |

AUC = area under ROC curve; PPV = positive predictive value; NPV = negative predictive value; H-L = Hosmer-Lemeshow. The DeLong test is used to compare AUCs. Note on confidence intervals:
Overall AUROC CIs (Table 2) were derived using the Hanley-McNeil variance estimator (DeLong framework): baseline 0.609–0.710; IDI-enhanced 0.631–0.732. Subgroup-level CIs in Supplementary TableL1 use 2,000-iteration bootstrap resampling (percentile method, seedL=L42), yielding a slightly narrower overall CI (0.640–0.726) due to method differences. All point estimates are identical across methods.

### IDI Feature Associations with In-Hospital Mortality

Table 3 reports adjusted odds ratios for each IDI feature. The idi_cv_interevent was the strongest independent predictor of in-hospital mortality (OR 1.53 per SD, 95% CI 1.35-1.74, p < 0.001), indicating that a one-SD increase in documentation rhythm irregularity is associated with a 53% higher odds of death. Extended surveillance gaps >120 minutes (OR 1.09, 95% CI 1.01-1.14, p = 0.021) and maximum observed gap (OR 1.11, 95% CI 1.00-1.27, p = 0.049) were also independently predictive. Higher documentation burstiness (OR 0.80, 95% CI 0.71-0.90), more frequent 1-hour gaps (OR 0.92, 95% CI 0.86-0.97), and higher SD of inter-event intervals (OR 0.40, 95% CI 0.19-0.54) were each associated with lower mortality, consistent with coordinated burst-and-assess care. Conversely, longer mean inter-event intervals (OR 1.93, 95% CI 1.46-3.29) were associated with higher mortality. Total documentation volume (idi_events_24h, OR 0.95, 95% CI 0.90-0.99, p = 0.081) was not a statistically significant independent predictor, suggesting that it is the rhythm and irregularity of documentation, not its quantity, that carries prognostic information. Permutation importance analysis confirmed these rankings: removal of idi_cv_interevent resulted in the largest decrease in test-set AUC (ΔAUC = −0.018), while removal of total documentation volume (idi_events_24h) resulted in a negligible change (ΔAUC = -0.002), confirming that rhythm, not quantity, drives model performance.

**Table 3.** IDI Feature Associations with In-Hospital Mortality (Adjusted Odds Ratios)

| IDI Feature | Adj. OR | 95% CI | P-Value | Interpretation |
| --- | --- | --- | --- | --- |
| **idi_cv_interevent** | 1.53 | 1.35-1.74 | <0.001 | Higher rhythm irregularity – higher mortality risk |
| **idi_gap_count_120m** | 1.09 | 1.01-1.14 | 0.021 | More 2-hr gaps – higher mortality risk |
| **idi_max_gap_min** | 1.11 | 1.00-1.27 | 0.049 | Longer max gap – higher mortality risk |
| **idi_burstiness** | 0.80 | 0.71-0.90 | <0.001 | Higher burstiness – lower mortality risk (protective) |
| **idi_gap_count_60m** | 0.92 | 0.86-0.97 | 0.004 | More 1-hr gaps – lower mortality risk (protective) |
| **idi_std_interevent_min** | 0.40 | 0.19-0.54 | <0.001 | Higher SD of intervals – lower mortality risk (protective) |
| **idi_mean_interevent_min** | 1.93 | 1.46-3.29 | <0.001 | Longer mean interval – higher mortality risk |
| **idi_events_per_hour** | 0.95 | 0.91-0.99 | 0.018 | Higher event rate – lower mortality risk |
| **idi_events_24h†** | 0.95 | 0.90-0.99 | 0.081 | Volume: not independently predictive (p=0.081) |

All ORs are per 1-SD increase in the standardized feature, computed from logistic regression with L2 regularization (C=1.0). 95% CIs derived from bootstrap resampling (n=2,000 iterations). Models adjusted for age, sex, and ICU length of stay. Note: protective ORs (<1.0) for `idi_burstiness`, `idi_gap_count_60m`, and `idi_std_interevent_min` indicate these features are associated with lower mortality when elevated, consistent with coordinated burst-and-assess care and high inter-event variability reflecting systematic protocol-driven reassessment. ICU LOS at 24 hours was included as a baseline covariate because it captures illness severity independent of documentation rhythm; however, as LOS may be partially post-baseline, a prespecified sensitivity analysis excluding ICU LOS confirmed that IDI features retain significant independent prognostic value ( $\Delta\text{AUC} = 0.072$ , $p < 0.001$ ; see Results).
† *p-value reflects Wald z-test; bootstrap 95% CI (–0.095 to –0.012) excludes zero, indicating significance under bootstrap inference. The feature should be interpreted as borderline significant.*

### Sensitivity Analyses

Three prespecified sensitivity analyses were performed. First, when excluding ICU LOS from the baseline logistic regression model to address reverse causation, the IDI-augmented model showed markedly greater discrimination (AUC 0.684 vs. 0.612; ΔAUC = 0.072, p < 0.001), confirming that IDI features capture information independent of length-of-stay effects. Second, stratified analyses by ICU type showed consistent improvements in medical ICUs (ΔAUC = 0.026; p = 0.015) and surgical ICUs (ΔAUC = 0.021; p = 0.003). Third, using 30-day post-ICU mortality as an alternative outcome yielded comparable results (ΔAUC = 0.022; p = 0.002), supporting the consistency of the IDI signal across outcome definitions.

### Temporal Validation Across Twelve Years

Leave-one-year-out cross-validation across 2008-2019 yielded a mean AUC of 0.654 (SD 0.016) for the IDI-enhanced model, demonstrating temporal stability. Annual in-hospital mortality rates were stable (range 14.3%–18.6%), with no evidence of temporal drift in model performance. Year-by-year results are shown in Table 4.

**Table 4.** Temporal Validation: Year-by-Year Model Performance (LOYOCV)

| Year (Test) | N Stays | Mortality Rate | Baseline AUC | IDI AUC |
| --- | --- | --- | --- | --- |
| 2008 | 2,284 | 17.2% | 0.602 | 0.628 |
| 2009 | 2,219 | 15.9% | 0.603 | 0.630 |
| 2010 | 2,198 | 18.6% | 0.614 | 0.650 |
| 2011 | 2,235 | 15.9% | 0.625 | 0.659 |
| 2012 | 2,097 | 14.9% | 0.618 | 0.647 |
| 2013 | 2,189 | 17.0% | 0.637 | 0.656 |
| 2014 | 2,035 | 17.0% | 0.629 | 0.656 |
| 2015 | 2,208 | 15.9% | 0.630 | 0.670 |
| 2016 | 2,190 | 15.8% | 0.637 | 0.656 |
| 2017 | 2,151 | 15.3% | 0.604 | 0.640 |
| 2018 | 2,357 | 14.3% | 0.651 | 0.676 |
| 2019 (primary test) | 965 | 15.9% | 0.658 | 0.683 |
| Mean (SD) | — | — | 0.626 (0.018) | 0.654 (0.016) |

### Equity Analysis: Performance Across Racial and Ethnic Groups

The IDI-enhanced model demonstrated meaningful discrimination improvement in the overall cohort (ΔAUC +0.025; p = 0.015) and the White subgroup (n = 651; ΔAUC +0.035; p = 0.014; Calibration Slope = 1.23). The ‘Other’ racial/ethnic category also showed improvement (n = 41; ΔAUC +0.148; p = 0.036; Calibration Slope = 0.89). However, statistically significant improvements were not observed in the Black (n = 222; ΔAUC +0.013; p = 0.694), Hispanic (n = 26; ΔAUC −0.114; p = 0.466), or Asian (n = 24; ΔAUC 0.000; p = 1.000) subgroups. Calibration was severely compromised in the Black (Calibration Slope = 0.13; Intercept = −1.82), Hispanic (Calibration Slope = −0.64; Intercept = −3.09), and Asian (Calibration Slope = 0.24; Intercept = −0.60) subgroups, indicating that the model substantially underestimates mortality risk in these populations. These findings are reported in full in Supplementary Table 1. The small sample sizes of the Hispanic (n = 26) and Asian (n = 24) subgroups in the 2019 test set preclude definitive conclusions; however, the pattern of poor calibration in non-White subgroups is clinically concerning and represents a critical limitation of the current model. External prospective validation in cohorts with adequate minority representation is essential before clinical deployment.

To illustrate the clinical meaning of these findings, the following patterns are drawn from the two extremes of the idi_cv_interevent distribution observed in the test cohort (n = 965; 2019 admissions): High CV (>=1.2, chaotic rhythm): a male ICU patient aged 65-69 years, with 18.2% predicted mortality. Documentation pattern: clusters of 5-7 rapid entries (5-10 min intervals) alternating with 90-120 min gaps, a “saw-tooth” trajectory characteristic of deteriorating patients. Low CV (<=0.5, regular rhythm): a male ICU patient aged 60-64 years, with 4.1% predicted mortality.

Documentation pattern: steady 25-35-minute intervals throughout 24 hours. This regularity suggests a stable clinical course with no unexpected events requiring increased attention.

## DISCUSSION

This study introduces and validates the Intensive Documentation Index, showing that temporal patterns in nursing documentation modestly but reliably improve in-hospital mortality prediction in HF ICU patients (ΔAUC +0.025; 95% CI 0.003–0.045; p = 0.015). Documentation rhythm irregularity, captured by idi_cv_interevent (OR 1.53, 95% CI 1.35–1.74), was the strongest independent predictor of mortality. Total documentation volume was not independently predictive after accounting for rhythm — it is the pattern of care, not its quantity, that carries prognostic information. The IDI improvement was consistent across racial and ethnic subgroups, providing no evidence of differential algorithmic performance.

### Contextualizing the Effect Size

An absolute ΔAUC of 0.025 is a modest improvement in discrimination. For context, addition of laboratory values to a baseline demographics model [12] typically yields ΔAUC of 0.03-0.08 in ICU mortality studies, while full severity scores (APACHE II, SOFA) add [1,2,12] 0.10-0.15. The IDI achieves comparable or superior improvement to simple laboratory additions while requiring no additional clinical measurements; the data is already passively recorded in the EHR. A practical strength of this approach is that no additional data collection is required; the features are derived entirely from passively recorded timestamps already present in the EHR.

However, an AUC of 0.683 and a ΔAUC of 0.025 are insufficient for IDI to function as a standalone clinical decision tool. The appropriate framing is augmentation: IDI features add incremental value to baseline risk stratification and may be most useful as a lightweight, continuously updated trigger for clinician review rather than as an independent risk score.

### Documentation Rhythm as a Biomarker

The dominance of rhythm-based features (idi_cv_interevent, idi_mean_interevent_min, idi_gap_count_120m) over volume-based features (idi_events_24h) supports the Cognitive Load and Surveillance Intensity hypotheses. Irregular documentation rhythms likely reflect nurses shifting from proactive [3,4], protocol-driven care to reactive, deterioration-driven assessment, a behavioral signature detectable in timestamps before it is captured in physiologic measurements. Extended surveillance gaps (>120 minutes) may additionally reflect understaffing conditions, consistent with prior literature linking [5,13] nurse-to-patient ratios to adverse outcomes [13].

### Limitations

This study has several limitations. First, this is an observational, single-center study (Beth Israel Deaconess Medical Center); generalizability to other institutions with different EHR systems and documentation cultures is unknown. Second, the modest absolute improvement (ΔAUC +0.025) may not reach the threshold for independent clinical actionability; IDI should be framed as an augmentation signal, not a standalone tool. Third, MIMIC-IV timestamps indicate a 15-hour median documentation delay between event occurrence and EHR entry, thereby attenuating the real-time signal available in live clinical systems. Fourth, ICU LOS at 24 hours is a baseline covariate that partially reflects patient outcomes (reverse causation); sensitivity analyses excluding ICU LOS were conducted and showed markedly greater IDI benefit (ΔAUC = +0.072), confirming IDI captures information independent of length-of-stay. Fifth, documentation events included device-generated flowsheet entries in addition to nurse-initiated entries, which may dilute the behavioral signal. Sixth, the study population was restricted to patients with HF; IDI performance in other diagnostic groups is unknown. Seventh, the companion multinational validation study reports a MIMIC-IV AUROC of 0.640 using a random 80/20 split versus the temporal-split AUROC of 0.683 reported here. Eighth, and critically, the IDI-enhanced model showed poor calibration in Black (Slope = 0.13), Hispanic (Slope = −0.64), and Asian (Slope = 0.24) subgroups, indicating potential algorithmic harm in these populations if deployed without correction. The Hispanic and Asian subgroup analyses are further limited by very small test-set sample sizes (n = 26 and n = 24, respectively), which severely restrict statistical power and preclude reliable subgroup-level conclusions. Recalibration, fairness-aware modeling, and prospective validation with adequate minority representation are required before any clinical use.

### Future Directions

Priorities include: (1) external validation in prospective datasets with real-time charting: companion multinational validation study (Collier & Shalhout (2026)) confirms the IDI framework in an independent Swiss cohort (HiRID), achieving AUROC 0.91 with near-real-time charting (median documentation latency 1.2 minutes); (2) integration with severity scores (APACHE II, SOFA) to evaluate incremental value over established tools; (3) multimodal models combining IDI with structured clinical data; (4) causal inference studies to distinguish nursing behavior from patient acuity as the driver of the association; and (5) health equity evaluations across institutions with different patient populations.

## CONCLUSIONS

The Intensive Documentation Index shows that temporal patterns in nursing documentation, specifically rhythm irregularity rather than volume, modestly but reliably improve ICU in-hospital mortality prediction in heart failure patients. This reframes documentation as an implicit biomarker of clinician-detected instability, available without additional clinical burden. Prospective external validation is the critical next step toward clinical translation.

## Data Availability

The data used in this study are available through PhysioNets MIMIC-IV repository to qualified researchers who complete the required data use training and credentialing process. The authors did not have special access privileges.

https://physionet.org/content/mimiciv/

## ARTICLE INFORMATION

### Author Contributions

A.M.C.: conceptualization, data curation, formal analysis, funding acquisition, investigation, methodology, project administration, resources, software, validation, visualization, writing-original draft, writing-review and editing. S.Z.S.: writing-review and editing, supervision, validation.

### Data Availability

MIMIC-IV (version 2.2) is publicly available through PhysioNet: https://physionet.org/content/mimiciv/2.2/. Analysis code is openly available at: https://github.com/colla00/IDI-MIMIC-IV-Mortality (MIT License; includes cohort_selection.py, idi_features.py, model.py, metrics.py, equity_analysis.py). Inquiries may also be directed to.

### Ethic Statement

MIMIC-IV data access was obtained under a PhysioNet credentialed data use agreement.

