## Supplemental Table 1 for "Development and Validation of the Intensive Documentation Index for ICU Mortality Prediction" (from another version of this preprint: version 4, 2026-03-20)

**Supplementary Table 1.** IDI-Enhanced Model Performance Across Racial and Ethnic Subgroups (Temporal Validation Test Set, 2019, n = 965)

| **Racial/Ethnic Subgroup** | **n (%)** | **Baseline AUC (95% CI)** | **IDI-Enhanced AUC (95% CI)** | **ΔAUC** | **p-value (DeLong)** | **Calibration Slope** | **Calibration Intercept** |
| --- | --- | --- | --- | --- | --- | --- | --- |
| **Overall** | **965 (100%)** | **0.658 (0.609–0.710)*** | **0.683 (0.640–0.726)*** | **+0.025** | **0.015** | **0.96** | **0.01** |
| **White** | **651 (67.5%)** | **0.620 (0.560–0.677)** | **0.655 (0.598–0.710)** | **+0.035** | **0.014** | **1.23** | **0.45** |
| **Black** | **222 (23.0%)** | **0.511 (0.361–0.658)** | **0.524 (0.392–0.659)** | **+0.013** | **0.694** | **0.13** | **−1.82** |
| **Hispanic** | **26 (2.7%)** | **0.523 (0.320–0.750)** | **0.409 (0.000–0.702)** | **−0.114** | **0.466** | **−0.64** | **−3.09** |
| **Asian** | **24 (2.5%)** | **0.500 (0.200–0.815)** | **0.500 (0.227–0.769)** | **0.000** | **1.000** | **0.24** | **−0.60** |
| **Other** | **41 (4.3%)** | **0.600 (0.405–0.805)** | **0.748 (0.579–0.905)** | **+0.148** | **0.036** | **0.89** | **−0.25** |

AUC = area under the receiver operating characteristic curve; CI = 95% confidence interval; ΔAUC = IDI-enhanced AUC minus baseline AUC; Cal. = calibration. Baseline model: age, sex, ICU length of stay. IDI-enhanced model: baseline + nine IDI temporal features (after leakage filter). Subgroup AUC confidence intervals derived from 2,000-iteration bootstrap resampling (percentile method, seed = 42). p-values from DeLong test for paired AUC comparison. Calibration slope and intercept estimated via logistic regression of outcome on logit-transformed predicted probabilities. Temporal validation test set: 2019 admissions only (n = 965). Subgroup-level n values reflect test-set composition; overall cohort racial composition reported in main text (Table 1).
